# Recent trends and geographic variation in delays for endoscopy and imaging in England: analysis of monthly diagnostics data

**DOI:** 10.1101/2025.11.06.25339656

**Authors:** Becky White, Matthew E Barclay, Georgios Lyratzopoulos, Ruth Swann, Igor Francetic, Matt Sutton, Gary Abel

## Abstract

**Background:** Timely diagnostic testing is essential for rapid diagnosis of potentially life-threatening conditions, but formal assessment of current trends and geographic variation in diagnostic delays in England is lacking.

**Methods:** We assessed recent trends in four imaging and four endoscopy tests in England using public data across 103 sub-Integrated Care Boards from 2019–2026. We compared the proportion of patients waiting ≥6 weeks in June 2026 versus June 2025 using mixed-effects binomial logistic regression. Changes in geographic variation (beyond that expected by chance) were quantified using random-effect variances. Local trends were examined using conditional estimates.

**Results:** Previous reductions in delays have stalled in the past year, even increasing for gastrointestinal endoscopies and MRI. From June 2025 to June 2026, delays increased for gastroscopy (24%–29%), colonoscopy (26%–31%), flexible sigmoidoscopy (28%–32%), and MRI (15%–23%). Substantial geographic variation persists, with three- to five-fold differences across the middle 50% of local areas. Local variation has increased significantly for MRI. Local trends diverged markedly, with delays falling by >50% in some areas and doubling in others.

**Conclusions:** Policy-makers should prioritise tackling deteriorating waiting times for gastrointestinal endoscopies and MRIs, and identifying local drivers of delays.

## Introduction

When patients present in primary care with symptoms of possible disease, timely use of relevant diagnostic tests is key to early diagnosis of life-threatening conditions such as cancer.

Rising demand and lasting disruption caused by the COVID-19 pandemic are thought to contribute to worsening delays in testing in England.^1^ According to NHS England’s target for 15 diagnostic tests, fewer than 1% of referred patients should wait 6 weeks or longer.^2^ This target has not been met since 2013. In January 2020, 4.4% of patients waited at least 6 weeks, but this rapidly deteriorated during the COVID pandemic to 58.5% in May 2020 and has remained well above 15% ever since.^2^

These nationwide performance statistics may mask substantial variation by local area. While relevant data is routinely published,^3^ reports have not formally quantified geographic variation in recent years. This has prevented the identification of localised bottlenecks in the health system, and understanding of the potential contribution of such bottlenecks to delayed cancer diagnosis and treatment.

Our research questions were:

1. What are the current national trends in imaging and endoscopy delays?
2. a) Is there geographic variation in delays beyond that caused by random chance, and b) is such variation currently widening or reducing?
3. Do current trends in delays vary by local area?

## Methods

### Data

We used data published by NHS England^3^ of the monthly number of patients waiting 6 weeks or longer for diagnostic tests. Data is reported for local areas, known previously as Clinical Commissioning Groups (CCGs) and recently as sub-Integrated Care Board locations (sub-ICBs), which we refer to as local areas. Mergers took place from 2020-21, reducing the number of areas from 197 in 2019 to 106 by 2025. We aggregated data for the areas that merged so that the same 103 units (current existing sub-ICBs) were followed throughout (3 sub-ICBs were excluded from analysis due to major boundary changes in June 2026).

We examined data for eight individual tests, comprising 4 endoscopic (gastroscopy, flexible sigmoidoscopy, colonoscopy, and cystoscopy) and 4 imaging (MRI, CT, non-obstetric ultrasound, echocardiography) investigations.

### Statistical analysis

To assess overall trends, we plotted the observed monthly proportion of patients waiting at least 6 weeks from January 2019 to June 2026, by local area (including the median local area and interquartile range).

For the rest of the analysis, we modelled local area data for two time points; the latest month (June 2026), compared to the same month in the previous year (June 2025). We did not model monthly observations continuously because of strong temporal autocorrelation within local areas and non-independence. This was caused by the dynamics of queue systems, where the size of a waiting list in a particular month will be directly determined by the waiting list in previous months, more so in areas with longer delays.^4,5^ Further, the outcome measure (patients waiting 6 weeks or longer), by definition incorporated patients from the previous month.

To minimise the influence of temporal dependence while accounting for seasonal effects, we used a repeated cross-sectional comparison of the latest calendar month with the same calendar month in the previous year. We assessed the stability of results by repeating the analysis for five alternative recent months (January, February, March, April, May).

For each diagnostic test studied, we fitted a mixed-effects binomial logistic regression model (Box 1), where the outcome was the monthly number of patients waiting 6 or more weeks as a proportion of all patients waiting at the end of that month. This included a random intercept for local area, which allowed the baseline proportion of patients waiting 6 weeks to vary by local area. The effect of year (June 2025 vs June 2026) was allowed to vary by local area.

#### Box 1. Mixed-effects binomial logistic regression model for each diagnostic test

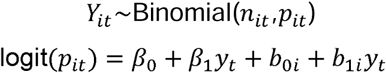

where:

- Y_it_ = number of patients waiting ≥6 weeks in local area i at time t
- n_it_ = total number of patients waiting
- *p_it_* = probability that a patient is waiting ≥6 weeks in local area i at time t
- β_0_ = Intercept
- *β*_1_*y_t_* = Fixed effect for year (June 2026 vs June 2025)
- *b_0i_* = Random effect for local area
- *b_1i_y_t_* = Local area specific random slope for year

To assess whether delays continued to improve in the past year (RQ 1), we extracted the fixed effect and p-value for year from the model.

To examine whether there was geographic variation in delays beyond that caused by random chance (RQ 2a), we used measures of variance between local areas from the model, which accounts for increased observed variance due to random chance, particularly in areas with small sample sizes (after Abel and Elliot).^6^ We extracted the modelled standard deviation for the local area random intercept in June 2026, which we used to calculate the 25^th^ and 75^th^ percentiles of the fitted distribution on a log-odds scale. We converted these back to proportions, giving the 50% midrange values that the majority of local areas lie within after accounting for chance, and the odds ratio difference between them. We compared this against the observed 50% midrange values calculated from the standard deviation of the observed local area values.

We also compared the modelled and observed normal distributions graphically. To do so, we transformed the modelled distribution from the log odds scale to proportion scale using the method outlined by Abel and Elliot.^6^ If true variation existed outside of random chance, this would be evident in the modelled 50% midrange values and graphical distributions, which would not be substantially smaller than the observed 50% midrange.

To examine whether variation was currently reducing (RQ 2b), we compared the modelled standard deviation for the local area random intercept for the most recent month (June 2026) with the comparison month (June 2025). We tested whether the difference in the standard deviation was statistically significant using bootstrapping (1,000 iterations per test).

To examine whether trends in delays vary by local area (RQ 3), we used the marginaleffects package to compare the predicted conditional proportion of patients waiting 6 weeks or longer in the current month (June 2026) and in a comparison month (June 2025). To conduct inference we used the delta method to estimate 95% confidence intervals around these proportions, and a Wald test to assess the null hypothesis that the proportion was the same in the current versus comparison month. Modelled proportions provided conservative estimates of delays in local areas with extreme values and small sample sizes, and ensured that intra-area differences between the two time periods could be formally compared.

### Software

Statistical analysis was conducted in R Studio version 4.5.2, using the following packages: lme4, marginaleffects, sf.

### Patient and public involvement

Patients were involved in the interpretation of this research and results discussion.

## Results

### What are the current national trends in imaging and endoscopy delays?

Overall, diagnostic test delays have reduced since the peak of the pandemic in 2020, but have not fully recovered (Figure 1, Appendix 1). Between June 2021 and June 2026, the observed national average proportion of patients waiting 6 weeks or longer fell for all endoscopy and two imaging tests studied: falling from 41% to 29% for gastroscopy, 41% to 30% for colonoscopy, 42% to 32% for flexible sigmoidoscopy, 35% to 26% for cystoscopy, 16% to 10% for CT, and 37% to 28% for echocardiography.

**Figure 1.**
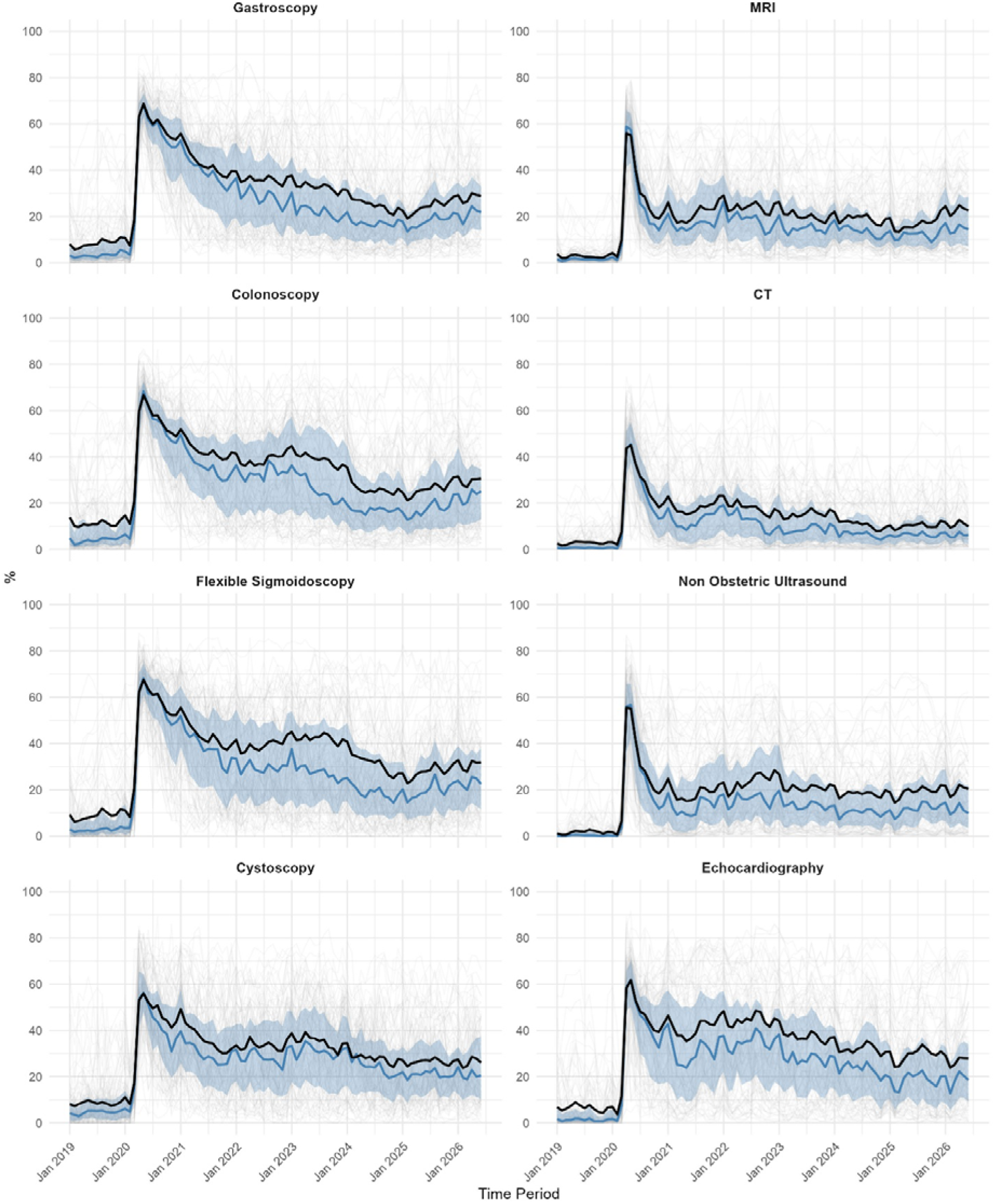
Percentage of patients waiting 6+ weeks for a test, by local area. National total (black), individual local areas (grey), median local area (blue), local area interquartile range (blue ribbon).

For MRI and non-obstetric ultrasound, the proportion of patients facing delays appeared to slightly increase from June 2022 to June 2026; 19% to 23% for MRI, and 15% to 21% for non-obstetric ultrasound.

In the past year, delays increased for three endoscopy tests. Observed proportions of patients waiting 6 weeks or longer increased from 24% in June 2025 to 29% in June 2026 (p = 0.003) for gastroscopy, 26% to 31% (p = 0.008) for colonoscopy, 28% to 32% (p = 0.043) for flexible sigmoidoscopy, and 15% to 23% (p = 0.002) for MRI. This was confirmed by model fixed effect odds ratios and 95% p-values (Table 1, Appendix 2). For cystoscopy, CT, non-obstetric ultrasound, and echocardiography, there were no statistically significant changes. In five sets of alternative models comparing other recent months (January, February, March, April, May) with their respective month in 2025, results were consistent; significant increases in delays for gastro-intestinal endoscopies and MRIs were also present in all comparisons (Appendix 3).

**Table 1.** Observed national proportions of patients waiting 6 weeks or longer for a diagnostic test, and modelled effects, June 2025 versus June 2026. *Fixed effect for year compares June 2026 with June 2025 from a mixed effects binomial logistic regression model

| Diagnostic test | National count (waiting ≥6 weeks) (2025-06) | National denominator (total waiting) (2025-06) | National % (waiting ≥6 weeks) (2025-06) | National denominator (total waiting) (2026-06) | National % (waiting ≥6 weeks) (2026-06) | Fixed effect for year (odds ratio) | P-value for fixed effect |
| --- | --- | --- | --- | --- | --- | --- | --- |
| Gastroscopy | 15326 | 63154 | 24.27 | 71027 | 28.91 | 1.294 | 0.003 |
| Colonoscopy | 16121 | 62186 | 25.92 | 73637 | 30.63 | 1.323 | 0.008 |
| Flexible Sigmoidoscopy | 5790 | 20670 | 28.01 | 22468 | 31.93 | 1.253 | 0.043 |
| Cystoscopy | 6261 | 23085 | 27.12 | 23685 | 26.07 | 0.95 | 0.566 |
| MRI | 48278 | 317363 | 15.21 | 380007 | 22.61 | 1.319 | 0.002 |
| CT | 17822 | 169460 | 10.52 | 192372 | 10.05 | 0.95 | 0.59 |
| Non Obstetric Ultrasound | 111559 | 587525 | 18.99 | 622717 | 20.52 | 0.923 | 0.539 |
| Echocardiography | 44523 | 147638 | 30.16 | 155749 | 27.9 | 1.009 | 0.949 |

### Is there geographic variation in delays beyond that caused by random chance?

In June 2026, depending on the diagnostic test, the odds of waiting 6 weeks or longer was three to five fold higher in the top of the middle range of local areas compared to the bottom (Table 2). For example, accounting for random chance, we estimated the true underlying 50% midrange of the proportion of patients waiting 6+ weeks for a gastroscopy in June 2026 across local areas to be 14% - 34%, corresponding to an odds ratio of 3.4.

**Table 2.**
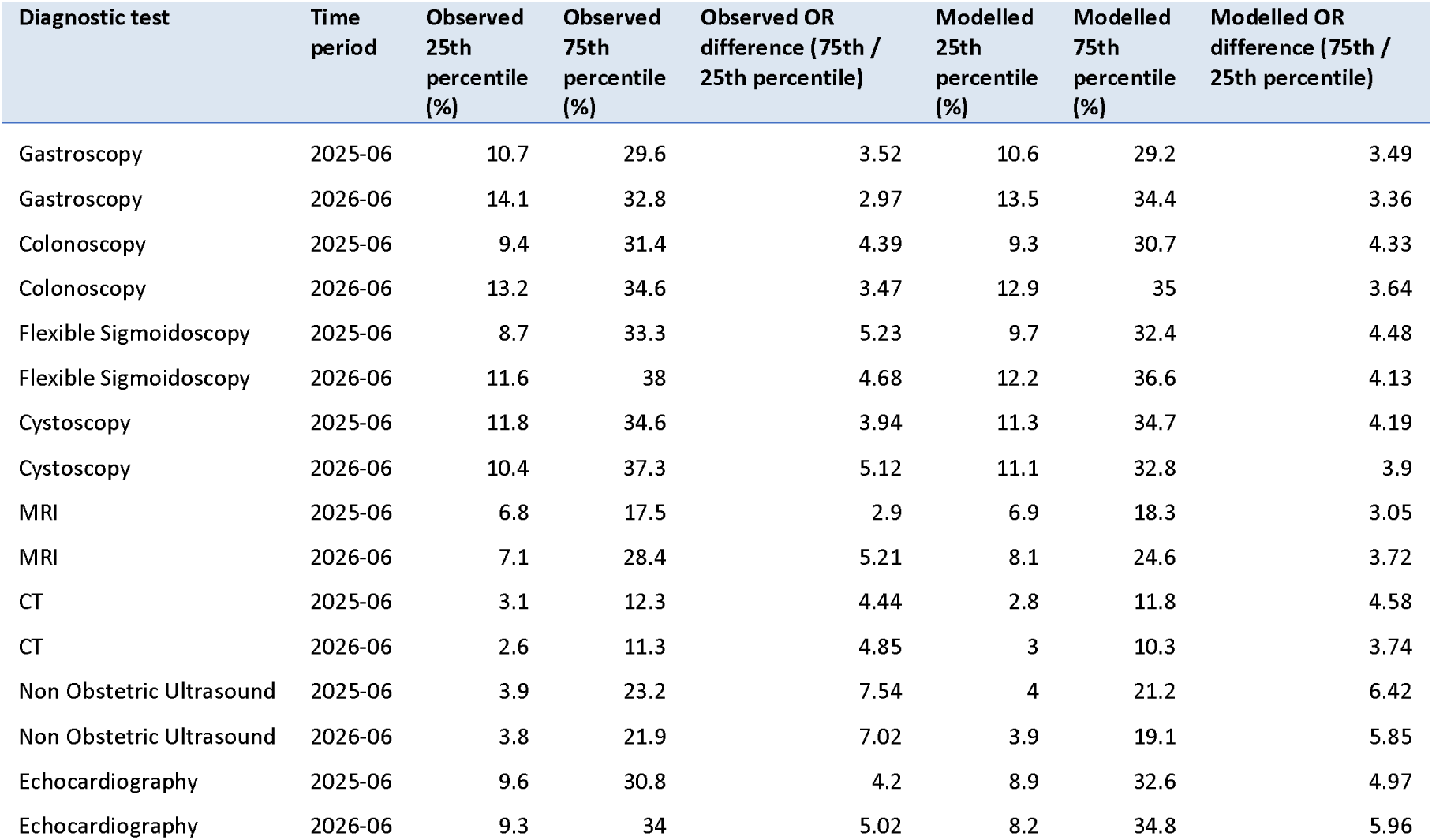
Observed and modelled percentile 25^th^ and 75^th^ (50% midrange) values by local area (% of patients waiting 6 weeks or longer). Underlying model coefficients are reported in Appendix 4. OR = odds ratio

| Diagnostic test | Time period | Observed 25th percentile (%) | Observed 75th percentile (%) | Observed OR difference (75th / 25th percentile) | Modelled 25th percentile (%) | Modelled 75th percentile (%) | Modelled OR difference (75th / 25th percentile) |
| --- | --- | --- | --- | --- | --- | --- | --- |
| Gastroscopy | 2025-06 | 10.7 | 29.6 | 3.52 | 10.6 | 29.2 | 3.49 |
| Gastroscopy | 2026-06 | 14.1 | 32.8 | 2.97 | 13.5 | 34.4 | 3.36 |
| Colonoscopy | 2025-06 | 9.4 | 31.4 | 4.39 | 9.3 | 30.7 | 4.33 |
| Colonoscopy | 2026-06 | 13.2 | 34.6 | 3.47 | 12.9 | 35 | 3.64 |
| Flexible Sigmoidoscopy | 2025-06 | 8.7 | 33.3 | 5.23 | 9.7 | 32.4 | 4.48 |
| Flexible Sigmoidoscopy | 2026-06 | 11.6 | 38 | 4.68 | 12.2 | 36.6 | 4.13 |
| Cystoscopy | 2025-06 | 11.8 | 34.6 | 3.94 | 11.3 | 34.7 | 4.19 |
| Cystoscopy | 2026-06 | 10.4 | 37.3 | 5.12 | 11.1 | 32.8 | 3.9 |
| MRI | 2025-06 | 6.8 | 17.5 | 2.9 | 6.9 | 18.3 | 3.05 |
| MRI | 2026-06 | 7.1 | 28.4 | 5.21 | 8.1 | 24.6 | 3.72 |
| CT | 2025-06 | 3.1 | 12.3 | 4.44 | 2.8 | 11.8 | 4.58 |
| CT | 2026-06 | 2.6 | 11.3 | 4.85 | 3 | 10.3 | 3.74 |
| Non Obstetric Ultrasound | 2025-06 | 3.9 | 23.2 | 7.54 | 4 | 21.2 | 6.42 |
| Non Obstetric Ultrasound | 2026-06 | 3.8 | 21.9 | 7.02 | 3.9 | 19.1 | 5.85 |
| Echocardiography | 2025-06 | 9.6 | 30.8 | 4.2 | 8.9 | 32.6 | 4.97 |
| Echocardiography | 2026-06 | 9.3 | 34 | 5.02 | 8.2 | 34.8 | 5.96 |

Little of the observed variation appeared to be caused by random chance, with the modelled and observed distributions in June 2026 being similar (Appendix 4 & Appendix 5). The midrange ratio was only slightly attenuated in modelled compared to observed results (Table 2).

### Is geographic variation currently reducing?

For all eight tests studied, there was no statistically significant difference between June 2026 and June 2025 in the size of variation between local areas in the proportion of patients experiencing delays, except for MRI, as shown by the modelled standard deviation for the local area random intercept (Appendix 6). For MRI, the standard deviation has increased from 0.83 on the log-odds scale in June 2025 to 0.97 in June 2026 (p = 0.0183).

In five alternative sets of models comparing other recent months (January, February, March, April, May) with their respective month in 2025, results were consistent; local area variation increased for MRI only, with the exception of an increase in variation for echocardiography in one model from March 2025 to March 2026 (Appendix 6).

### Do trends in delays vary by local area?

The proportion of patients facing delays within individual local areas was often very different in June 2026 than in 2025, with changes in local areas substantially differing in direction and scale (Figure 2). For instance, for gastroscopy, the modelled proportion of patients waiting at least 6 weeks reduced by at least half in 5 local areas, while it doubled or more in 21 local areas (Appendix 7 & Appendix 8). Annual changes in performance did not show obvious geographical patterns, with areas often improving for one test but getting worse for another (Figure 2).

**Figure 2.**
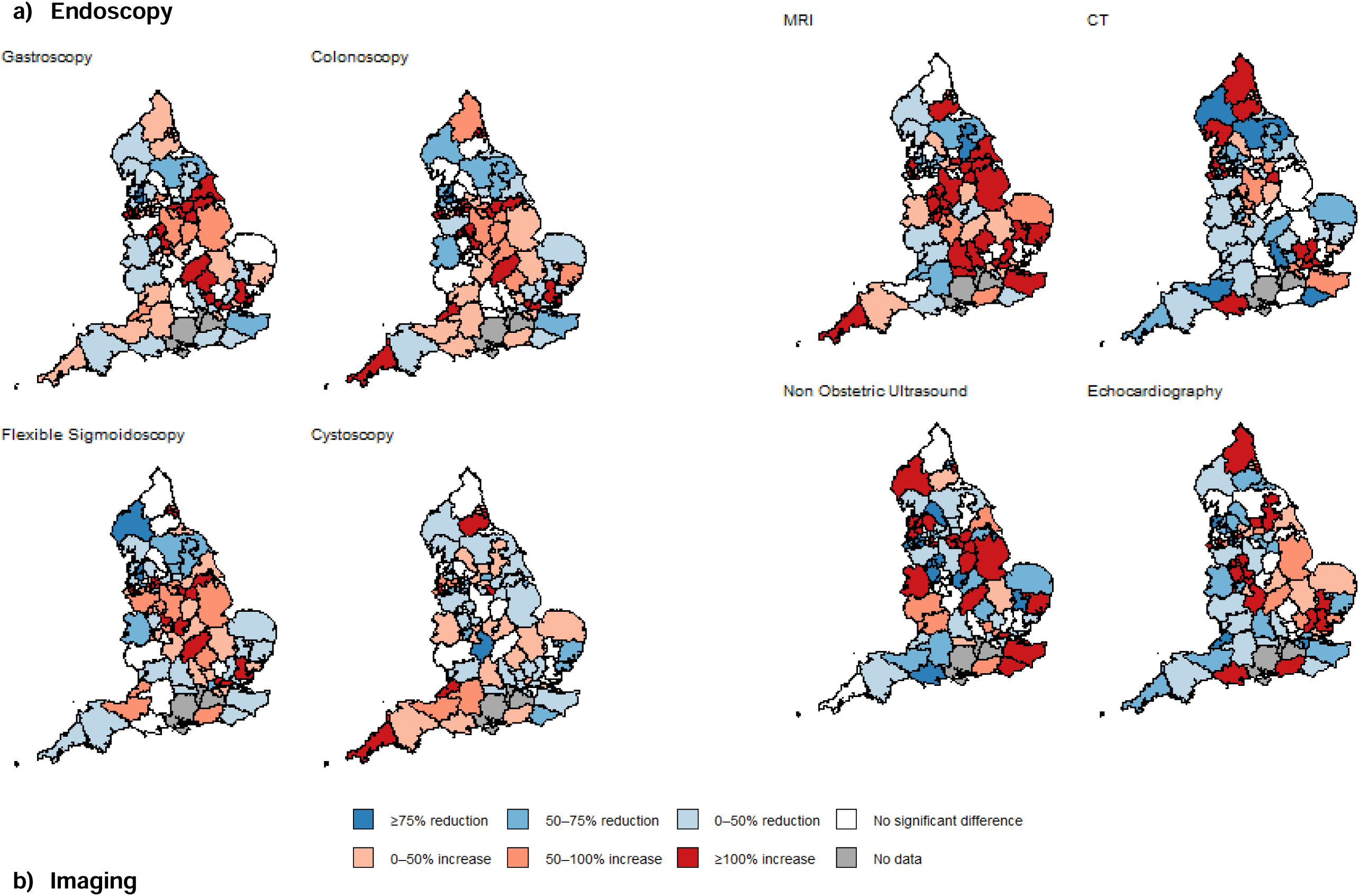
Relative change from June 2025 to June 2026 in the proportion of patients waiting 6 weeks or longer for a test, by local area.

Overall, local areas with higher proportions of patients waiting 6 weeks or longer in June 2025 tended to decrease (i.e. improve) the most by June 2026 (Appendix 9). This was also indicated by modelled correlation of random effects, which was negative for all tests studied (Appendix 2).

## Discussion

### Main finding of this study

Recent improvements in delays for imaging and endoscopy investigations have stalled in the past year, and even begun to reverse for gastro-intestinal endoscopies and MRI.

There is substantial variation between local areas in the scale of delays for all the tests studied, beyond that caused by random chance. Such variation is persistent over the past year, and relative differences have widened for MRI. Overall variation masks substantial differences in the direction and scale of change in delays within local areas; with the proportion of patients facing delays reducing by over 75% in some areas and doubling in others.

### What is already known on this topic and what this study adds

#### Current national trends

Our analysis contributes a rigorous and up to date assessment of trends in diagnostic delays in England. The initial COVID-related disruption to diagnostic services in England, and their partial recovery, were regularly tracked by both the BMA and Nuffield Trust; but these are no longer updated.^7,8^ The recent stalling in recovery that we highlight concords with a UK Parliament report, based on data up to March 2025.^9^ Our findings regarding national trends are reflected in statistical reports accompanying the NHS England dataset, however, these reports do not formally test for statistical differences in the national trend in the past year.^10^

There is no known study quantifying recent diagnostic test waiting times within subnational geographies of comparable countries. However, across OECD countries, post-COVID backlogs in diagnostic endoscopies and imaging,^11^ and persistent delays for elective surgeries have been documented.^12^

Persistent delays in diagnostic testing in England have been attributed to rising demand since before the pandemic, particularly for cancer diagnostic tests, which has not been matched by increased supply of equipment and workforce. For imaging, there is a lack of CT and MRI scanners per head compared to European counterparts, though there is no equivalent data for endoscopy facilities.^1^ What is unclear is why post-pandemic recovery in diagnostic waiting times has recently stalled. A recent UK parliament committee report suggests a number of failings, including the absence of a target to reduce waiting times for diagnostic tests in NHS England’s 2025-26 operational planning guidance, and an unclear evaluation plan for its Community Diagnostic Centres (CDCs) programme.^9^

#### Geographic variation in delays and divergent local trends

Diagnostic waiting times data in England by local area has been publicly available for some time, and accompanying reports describe geographic variation in delays, but this is only presented for broad region and all tests combined.^10^ Our study is the first to formally quantify the scale of local variation in delays for each test, and recent changes.

It is easy to overlook the substantial volatility over time within each local area. Our study is the first to compare area-specific trends in diagnostic delays in England, and proposes a solid methodological framework to do so. A previous evaluation of Community Diagnostic Centre (CDCs) using the same dataset from 2018 to 2023 also noted differences between hospital Trusts in the magnitude and trend in delays for diagnostic tests.^13^

We are not aware of research into the specific processes driving changes in delays for tests in different local areas in England, although various general theories have been proposed. Local demand may be affected by population need, long-term impacts of local COVID lockdowns (i.e. increased incidence of preventable diseases),^14^ local awareness campaigns,^15^ GP referral practices,^16^ screening,^17^ primary care-based triage tests (e.g.

Faecal Occult Immunochemical Testing (FIT) for patients with colorectal cancer symptoms, or following its future roll-out, Cytosponge for oesophageal cancer), or in theory, repeat testing caused by miscommunication between healthcare providers or invalid test results. Supply might be affected by scanner availability, efficiency or staffing. New service configurations can have complex effects; Community Diagnostic Centres could enhance capacity or, counterintuitively, stimulate demand.^13^ In addition, delays can be caused by temporary mismatches between supply and demand, when there is no flexibility to quickly increase supply to accommodate sudden influxes in demand.^18^

Finally, a novel finding of this study is the negative correlation between area-level waiting times from one year to the next, after accounting for chance. Our findings suggest some form of external regulation of the system. Poor-performing areas often improve, while areas that are performing well often become worse. It may be that special measures are implemented to bring excessive waiting lists under control, but the deterioration of high-performing areas also suggest some form of self-regulation where patients or GPs reduce demands for tests in the face of excessive delays – and then rapidly increase demands when delays reduce.^19^

### Limitations of this study

Our modelling approach compared geographic variation over time, while accounting for chance variation. We did not model monthly observations continuously because of strong temporal autocorrelation within local areas, which, if not explicitly modelled, could bias estimates of between-area variance. Time series modelling approaches explicitly account for temporal autocorrelation (e.g. ARIMA-type models),^20^ but are primarily suited to forecasting or intervention analysis, and do not naturally extend to estimating between-area variation across many units. Our sensitivity analysis indicated that the choice of comparison months is unlikely to affect the overall results. Future access to patient-level data could support more sophisticated longitudinal analyses by allowing individual patients to be followed across consecutive months and waiting times to be estimated directly.

The data we used is based on waiting times for the local commissioning area of the patient’s GP practice. Patients could travel to an NHS hospital Trust geographically located either within their GP practice’s local commissioning area or a neighbouring area. This means that delays in one commissioning area could partly reflect delays at hospital Trusts in a neighbouring area. This may introduce spatial correlation between neighbouring areas that was not explicitly modelled. In addition, changes were made to geographic boundaries in four areas in 2022, which we did not account for in our analysis.^21^

The data include tests conducted for any reason, including diagnosing non-life-threatening conditions, and subsequent follow up tests following screening, where delays over 6 weeks may not be as consequential. Quantification of delays is needed specifically when time-sensitive and potentially life-threatening disease (e.g. cancer) is suspected. Some local hospitals may prioritise these patients and others may not, which could exaggerate variation in delays for these patients.

## Conclusions

In summary, partial improvements in diagnostic waiting times in England have stalled in the past year, and even partially reversed for gastro-intestinal endoscopies and MRIs.

Sizeable geographic variation in delays persists and is increasing for MRIs. Local areas experience large differences in the scale and direction of changes in delays. Improvement efforts should prioritise reducing waiting times nationwide, while investigating the causes of volatility in local diagnostic delays, including local diagnostic capacity and service configuration, and demand.

## Author contributions

BW conceived the study. BW, MB, GL and GA designed the study. BW managed and analysed the data, and RS quality assured data analysis. BW drafted the manuscript. MB and GA advised on statistical methods. MS, IF, and RS advised on interpretation and discussion. All authors contributed to interpretation and revisions to the manuscript.

## Acknowledgements

This study is based on data published by NHS England. Dr Arman Motesharei downloaded the publicly available data. We would like to thank patient and public representatives who contributed to the results discussion, namely Julie Callin, Cathy Mullins, Clara Martins de Barros, and David Holden, also Evelyn Brunsdon for co-ordinating patient and public engagement.

## Funding

This work was supported by a Cancer Research UK International Alliance for Cancer Early Detection (ACED) Pathway Award (EDDAPA-2024/100015) granted to BW. This project is also funded by the NIHR Policy Research Programme (Policy Research Unit on Cancer Awareness, Screening and Early Diagnosis, reference PR-PRU-NIHR206132). The views expressed are those of the authors and not necessarily those of the NIHR or the Department of Health and Social Care.

## Conflict of interest

All authors declare no competing interests.

## Ethics approval

Not required.

## Data availability

Data are publicly available from https://www.england.nhs.uk/statistics/statistical-work-areas/diagnostics-waiting-times-and-activity. A fully compiled and processed dataset, and all analytical code, is available at https://github.com/rmjlrwh/DxWaitingTimes2.

## Notes

### Competing Interest Statement

The authors have declared no competing interest.

### Author Declarations

Data are publicly available from https://www.england.nhs.uk/statistics/statistical-work-areas/diagnostics-waiting-times-and-activity.

### Summary of Updates

Analysis updated in August 2026 to include data up to June 2026, and reformatted for a new journal submission.

