## Appendix for "Recent trends and geographic variation in delays for endoscopy and imaging in England: analysis of monthly diagnostics data"

**Appendices**

**Appendix 1. Observed national proportions of patients waiting 6 weeks or longer for a diagnostic test, January 2019 to June 2026**

*Preview: Full appendix available at* <https://github.com/rmjlrwh/DxWaitingTimes2>

| **Diagnostic test** | **Year-Month** | **National count (waiting ≥6 weeks)** | **National denominator (total waiting)** | **National % (waiting ≥6 weeks)** | **Median local area % (waiting ≥6 weeks)** | **25th percentile local area %** | **75th percentile local area %** | **N local areas** |
| --- | --- | --- | --- | --- | --- | --- | --- | --- |
| Gastroscopy | 2019-01 | 3334 | 41637 | 8.01 | 3.14 | 1.39 | 7.05 | 103 |
| Gastroscopy | 2019-02 | 2474 | 42936 | 5.76 | 2.15 | 0.93 | 5 | 103 |
| Gastroscopy | 2019-03 | 2609 | 42853 | 6.09 | 2.55 | 1.07 | 5.09 | 103 |
| Gastroscopy | 2019-04 | 6410 | 86652 | 7.4 | 3.12 | 1.48 | 7.08 | 103 |
| Gastroscopy | 2019-05 | 3345 | 43825 | 7.63 | 2.99 | 1.32 | 7.88 | 103 |
| Gastroscopy | 2019-06 | 3370 | 43095 | 7.82 | 2.83 | 0.98 | 7.01 | 103 |
| Gastroscopy | 2019-07 | 3425 | 42787 | 8 | 2.33 | 1.23 | 6.8 | 103 |
| Gastroscopy | 2019-08 | 4292 | 41936 | 10.23 | 3.64 | 1.73 | 9.47 | 103 |
| Gastroscopy | 2019-09 | 3995 | 42552 | 9.39 | 3.66 | 1.28 | 9.55 | 103 |
| Gastroscopy | 2019-10 | 4095 | 46200 | 8.86 | 3.37 | 1.37 | 8.18 | 103 |
| Gastroscopy | 2019-11 | 4199 | 46351 | 9.06 | 3.45 | 1.63 | 8.6 | 103 |

**Appendix 2. Full model terms for main model (June 2026 vs June 2025)**

| **Diagnostic test** | **Fixed effect for year (log-odds)** | **Fixed effect for year (odds ratio)** | **P-value for fixed effect** | **Random effect variance: intercept** | **Random effect variance: time slope** | **Covariance between random intercept and time slope** | **Correlation between random intercept and time slope** |
| --- | --- | --- | --- | --- | --- | --- | --- |
| Gastroscopy | 0.258 | 1.294 | 0.003 | 0.86 | 0.73 | -0.393 | -0.495 |
| Colonoscopy | 0.28 | 1.323 | 0.008 | 1.179 | 1.086 | -0.674 | -0.596 |
| Flexible Sigmoidoscopy | 0.226 | 1.253 | 0.043 | 1.236 | 1.092 | -0.611 | -0.526 |
| Cystoscopy | -0.051 | 0.95 | 0.566 | 1.129 | 0.672 | -0.39 | -0.448 |
| MRI | 0.277 | 1.319 | 0.002 | 0.683 | 0.829 | -0.282 | -0.375 |
| CT | -0.051 | 0.95 | 0.59 | 1.27 | 0.859 | -0.585 | -0.56 |
| Non Obstetric Ultrasound | -0.08 | 0.923 | 0.539 | 1.9 | 1.728 | -0.956 | -0.527 |
| Echocardiography | 0.009 | 1.009 | 0.949 | 1.415 | 1.821 | -0.743 | -0.463 |

**Appendix 3. Sensitivity analysis comparing national trends for main models (June 2026 vs June 2025) with five sets of alternative cross-sectional models (May, April, March, February, January)**

| **Diagnostic test** | **Model** | **Fixed effect for year (odds ratio)** | **P-value for fixed effect** |
| --- | --- | --- | --- |
| Gastroscopy | Jun 2026 vs Jun 2025 | 1.294 | 0.003 |
| Gastroscopy | May 2026 vs May 2025 | 1.396 | 0 |
| Gastroscopy | Apr 2026 vs Apr 2025 | 1.552 | 0 |
| Gastroscopy | Mar 2026 vs Mar 2025 | 1.429 | 0 |
| Gastroscopy | Feb 2026 vs Feb 2025 | 1.535 | 0 |
| Gastroscopy | Jan 2026 vs Jan 2025 | 1.367 | 0 |
| Colonoscopy | Jun 2026 vs Jun 2025 | 1.323 | 0.008 |
| Colonoscopy | May 2026 vs May 2025 | 1.382 | 0.001 |
| Colonoscopy | Apr 2026 vs Apr 2025 | 1.382 | 0.002 |
| Colonoscopy | Mar 2026 vs Mar 2025 | 1.427 | 0 |
| Colonoscopy | Feb 2026 vs Feb 2025 | 1.574 | 0 |
| Colonoscopy | Jan 2026 vs Jan 2025 | 1.44 | 0 |
| Flexible Sigmoidoscopy | Jun 2026 vs Jun 2025 | 1.253 | 0.043 |
| Flexible Sigmoidoscopy | May 2026 vs May 2025 | 1.35 | 0.011 |
| Flexible Sigmoidoscopy | Apr 2026 vs Apr 2025 | 1.483 | 0 |
| Flexible Sigmoidoscopy | Mar 2026 vs Mar 2025 | 1.494 | 0 |
| Flexible Sigmoidoscopy | Feb 2026 vs Feb 2025 | 1.471 | 0.001 |
| Flexible Sigmoidoscopy | Jan 2026 vs Jan 2025 | 1.299 | 0.015 |
| Cystoscopy | Jun 2026 vs Jun 2025 | 0.95 | 0.566 |
| Cystoscopy | May 2026 vs May 2025 | 1.054 | 0.565 |
| Cystoscopy | Apr 2026 vs Apr 2025 | 1.057 | 0.526 |
| Cystoscopy | Mar 2026 vs Mar 2025 | 0.917 | 0.31 |
| Cystoscopy | Feb 2026 vs Feb 2025 | 1.051 | 0.577 |
| Cystoscopy | Jan 2026 vs Jan 2025 | 1.044 | 0.658 |
| MRI | Jun 2026 vs Jun 2025 | 1.319 | 0.002 |
| MRI | May 2026 vs May 2025 | 1.264 | 0.01 |
| MRI | Apr 2026 vs Apr 2025 | 1.412 | 0 |
| MRI | Mar 2026 vs Mar 2025 | 1.45 | 0 |
| MRI | Feb 2026 vs Feb 2025 | 1.348 | 0.002 |
| MRI | Jan 2026 vs Jan 2025 | 1.185 | 0.112 |
| CT | Jun 2026 vs Jun 2025 | 0.95 | 0.59 |
| CT | May 2026 vs May 2025 | 0.928 | 0.45 |
| CT | Apr 2026 vs Apr 2025 | 1.09 | 0.377 |
| CT | Mar 2026 vs Mar 2025 | 1.036 | 0.737 |
| CT | Feb 2026 vs Feb 2025 | 1.037 | 0.706 |
| CT | Jan 2026 vs Jan 2025 | 1.157 | 0.114 |
| Non Obstetric Ultrasound | Jun 2026 vs Jun 2025 | 0.923 | 0.539 |
| Non Obstetric Ultrasound | May 2026 vs May 2025 | 0.871 | 0.283 |
| Non Obstetric Ultrasound | Apr 2026 vs Apr 2025 | 1.06 | 0.627 |
| Non Obstetric Ultrasound | Mar 2026 vs Mar 2025 | 1.173 | 0.174 |
| Non Obstetric Ultrasound | Feb 2026 vs Feb 2025 | 1.162 | 0.175 |
| Non Obstetric Ultrasound | Jan 2026 vs Jan 2025 | 1.132 | 0.238 |
| Echocardiography | Jun 2026 vs Jun 2025 | 1.009 | 0.949 |
| Echocardiography | May 2026 vs May 2025 | 0.993 | 0.949 |
| Echocardiography | Apr 2026 vs Apr 2025 | 1.173 | 0.108 |
| Echocardiography | Mar 2026 vs Mar 2025 | 1.079 | 0.456 |
| Echocardiography | Feb 2026 vs Feb 2025 | 1.021 | 0.847 |
| Echocardiography | Jan 2026 vs Jan 2025 | 0.909 | 0.3 |

**Appendix 4. Terms used to calculate modelled 50% midrange values – main model**

| **Diagnostic test** | **Mean (log-odds scale) — Jun 2025** | **Variance (log-odds scale) — Jun 2025** | **SD (log-odds scale) — Jun 2025** | **25th percentile (%) — Jun 2025** | **75th percentile (%) — Jun 2025** | **Mean (log-odds scale) — Jun 2026** | **Variance (log-odds scale) — Jun 2026** | **SD (log-odds scale) — Jun 2026** | **Difference in SD (latest minus previous, log-odds scale) — Jun 2026** | **SD difference lower bound (95% CI) — Jun 2026** | **SD difference upper bound (95% CI) — Jun 2026** | **P-value for change in SD — Jun 2026** | **25th percentile (%) — Jun 2026** | **75th percentile (%) — Jun 2026** |
| --- | --- | --- | --- | --- | --- | --- | --- | --- | --- | --- | --- | --- | --- | --- |
| Gastroscopy | -1.51 | 0.86 | 0.93 | 10.56 | 29.21 | -1.25 | 0.81 | 0.9 | -0.0299 | -0.1607 | 0.1008 | 0.6536 | 13.49 | 34.36 |
| Colonoscopy | -1.54 | 1.18 | 1.09 | 9.3 | 30.74 | -1.26 | 0.92 | 0.96 | -0.1285 | -0.2684 | 0.0114 | 0.0717 | 12.89 | 35.01 |
| Flexible Sigmoidoscopy | -1.49 | 1.24 | 1.11 | 9.66 | 32.4 | -1.26 | 1.11 | 1.05 | -0.0601 | -0.2575 | 0.1372 | 0.5503 | 12.25 | 36.58 |
| Cystoscopy | -1.35 | 1.13 | 1.06 | 11.26 | 34.73 | -1.4 | 1.02 | 1.01 | -0.0527 | -0.193 | 0.0876 | 0.4617 | 11.11 | 32.79 |
| MRI | -2.05 | 0.68 | 0.83 | 6.86 | 18.34 | -1.77 | 0.95 | 0.97 | 0.147 | 0.0248 | 0.2691 | 0.0183 | 8.09 | 24.65 |
| CT | -2.78 | 1.27 | 1.13 | 2.83 | 11.76 | -2.83 | 0.96 | 0.98 | -0.1475 | -0.3167 | 0.0217 | 0.0876 | 2.97 | 10.28 |
| Non Obstetric Ultrasound | -2.24 | 1.9 | 1.38 | 4.02 | 21.18 | -2.32 | 1.72 | 1.31 | -0.0681 | -0.2743 | 0.1382 | 0.5177 | 3.89 | 19.15 |
| Echocardiography | -1.53 | 1.41 | 1.19 | 8.86 | 32.59 | -1.52 | 1.75 | 1.32 | 0.1335 | -0.0702 | 0.3372 | 0.1989 | 8.22 | 34.8 |

**Appendix 5. Observed and modelled distributions of variation by local area in the proportion of patients waiting 6 weeks or longer for a test – June 2026**

a) Endoscopy

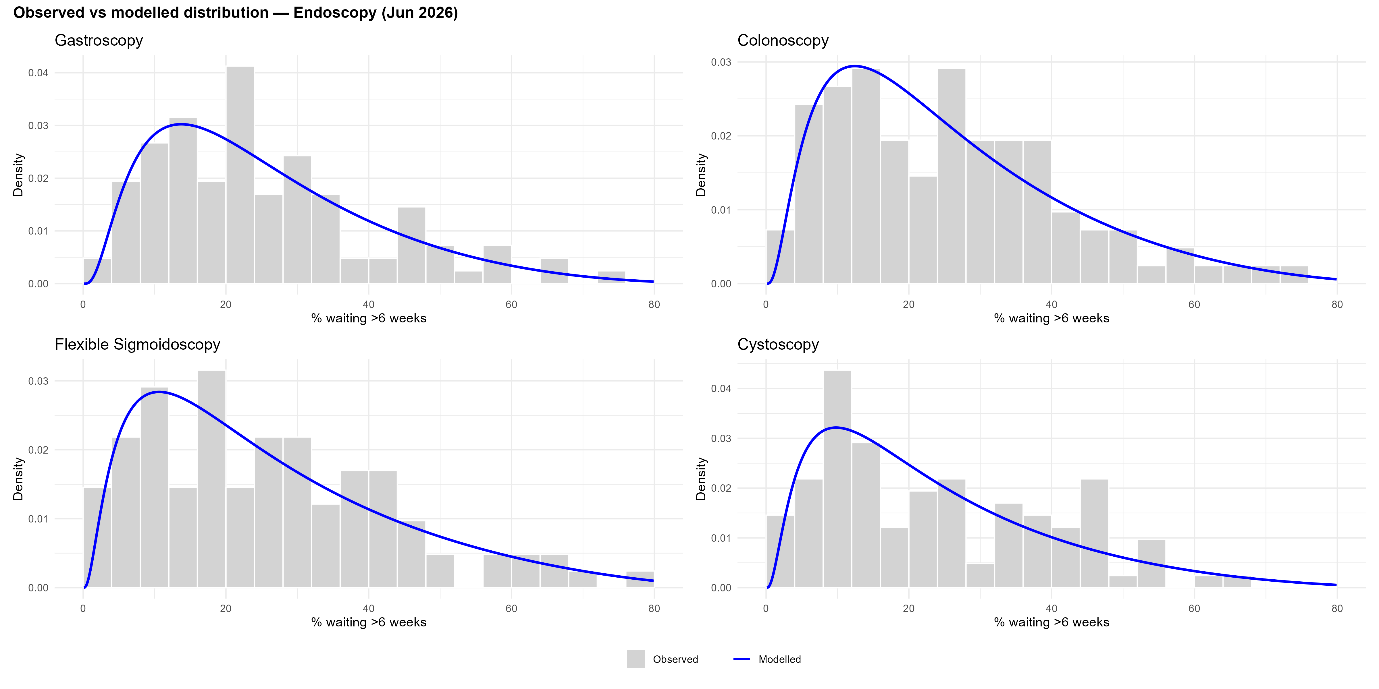

b) Imaging

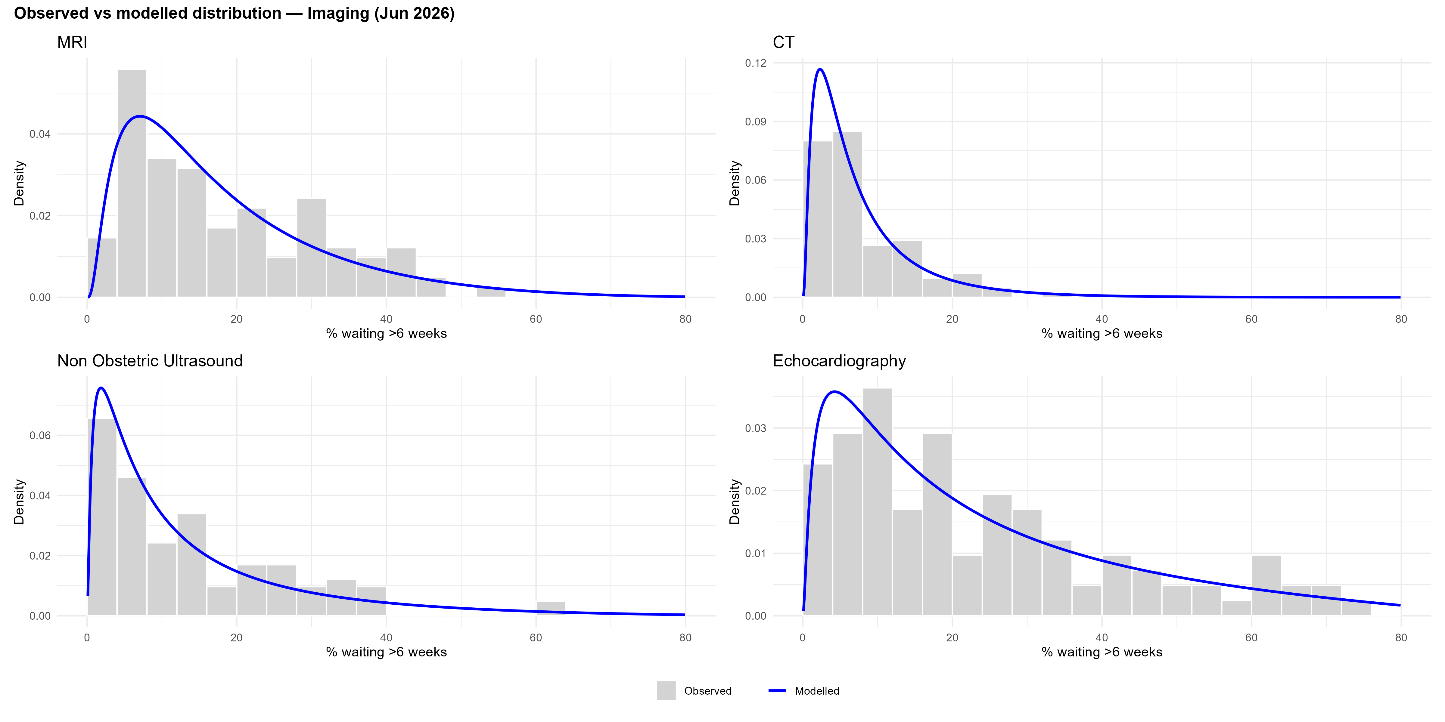

**Appendix 6. Modelled variation between local areas in the proportion of patients waiting 6 weeks or longer for a test, sensitivity analysis comparing main models (June 2026 vs June 2025) with five sets of alternative cross-sectional models (May, April, March, February, January)**

*SD: Standard deviation of the local area random intercept. Bootstrapped confidence intervals & p-values calculated from the standard error of differences in the standard deviation in month x 2025 vs month x 2026, generated from 1,000 model iterations

| **Diagnostic test** | **Model** | **SD (log-odds scale) — Month x, 2025** | **SD (log-odds scale) — Month x, 2026** | **Difference in SD (latest minus previous, log-odds scale)** | **SD difference lower bound (95% CI)** | **SD difference upper bound (95% CI)** | **P-value for change in SD** |
| --- | --- | --- | --- | --- | --- | --- | --- |
| Gastroscopy | Jun 2026 vs Jun 2025 | 0.93 | 0.9 | -0.0299 | -0.1607 | 0.1008 | 0.6536 |
| Gastroscopy | May 2026 vs May 2025 | 0.93 | 0.92 | -0.0059 | -0.1421 | 0.1303 | 0.932 |
| Gastroscopy | Apr 2026 vs Apr 2025 | 0.87 | 0.93 | 0.0571 | -0.083 | 0.1971 | 0.4244 |
| Gastroscopy | Mar 2026 vs Mar 2025 | 0.89 | 0.89 | 0.0056 | -0.1462 | 0.1574 | 0.942 |
| Gastroscopy | Feb 2026 vs Feb 2025 | 0.97 | 0.9 | -0.0713 | -0.2323 | 0.0898 | 0.3857 |
| Gastroscopy | Jan 2026 vs Jan 2026 | 0.91 | 0.89 | -0.0193 | -0.1967 | 0.158 | 0.8307 |
| Colonoscopy | Jun 2026 vs Jun 2025 | 1.09 | 0.96 | -0.1285 | -0.2684 | 0.0114 | 0.0717 |
| Colonoscopy | May 2026 vs May 2025 | 1.11 | 1.01 | -0.0992 | -0.245 | 0.0466 | 0.1825 |
| Colonoscopy | Apr 2026 vs Apr 2025 | 1.04 | 1.02 | -0.0214 | -0.1697 | 0.127 | 0.7778 |
| Colonoscopy | Mar 2026 vs Mar 2025 | 1.09 | 1.05 | -0.0346 | -0.2054 | 0.1362 | 0.691 |
| Colonoscopy | Feb 2026 vs Feb 2025 | 1.1 | 1.06 | -0.0481 | -0.2028 | 0.1066 | 0.5423 |
| Colonoscopy | Jan 2026 vs Jan 2026 | 1.05 | 1.06 | 0.0039 | -0.1439 | 0.1517 | 0.9587 |
| Flexible Sigmoidoscopy | Jun 2026 vs Jun 2025 | 1.11 | 1.05 | -0.0601 | -0.2575 | 0.1372 | 0.5503 |
| Flexible Sigmoidoscopy | May 2026 vs May 2025 | 1.14 | 0.96 | -0.1837 | -0.3754 | 0.0079 | 0.0602 |
| Flexible Sigmoidoscopy | Apr 2026 vs Apr 2025 | 1.02 | 0.95 | -0.0694 | -0.243 | 0.1041 | 0.4329 |
| Flexible Sigmoidoscopy | Mar 2026 vs Mar 2025 | 1.1 | 0.98 | -0.1252 | -0.3416 | 0.0911 | 0.2565 |
| Flexible Sigmoidoscopy | Feb 2026 vs Feb 2025 | 1.07 | 1.11 | 0.0379 | -0.1784 | 0.2541 | 0.7316 |
| Flexible Sigmoidoscopy | Jan 2026 vs Jan 2026 | 1.03 | 1.07 | 0.0455 | -0.149 | 0.24 | 0.6468 |
| Cystoscopy | Jun 2026 vs Jun 2025 | 1.06 | 1.01 | -0.0527 | -0.193 | 0.0876 | 0.4617 |
| Cystoscopy | May 2026 vs May 2025 | 1.07 | 1.02 | -0.0539 | -0.2142 | 0.1065 | 0.5103 |
| Cystoscopy | Apr 2026 vs Apr 2025 | 0.98 | 0.96 | -0.0207 | -0.1797 | 0.1382 | 0.798 |
| Cystoscopy | Mar 2026 vs Mar 2025 | 0.93 | 0.98 | 0.055 | -0.1077 | 0.2177 | 0.5076 |
| Cystoscopy | Feb 2026 vs Feb 2025 | 1.06 | 0.96 | -0.0995 | -0.2408 | 0.0418 | 0.1677 |
| Cystoscopy | Jan 2026 vs Jan 2026 | 1 | 0.94 | -0.0613 | -0.1937 | 0.0711 | 0.3644 |
| MRI | Jun 2026 vs Jun 2025 | 0.83 | 0.97 | 0.147 | 0.0248 | 0.2691 | 0.0183 |
| MRI | May 2026 vs May 2025 | 0.8 | 0.98 | 0.1725 | 0.041 | 0.304 | 0.0101 |
| MRI | Apr 2026 vs Apr 2025 | 0.83 | 0.96 | 0.1374 | 0.005 | 0.2697 | 0.0419 |
| MRI | Mar 2026 vs Mar 2025 | 0.83 | 0.99 | 0.1614 | 0.0393 | 0.2835 | 0.0096 |
| MRI | Feb 2026 vs Feb 2025 | 0.84 | 1 | 0.1549 | 0.0223 | 0.2875 | 0.022 |
| MRI | Jan 2026 vs Jan 2026 | 0.77 | 0.98 | 0.2047 | 0.0565 | 0.353 | 0.0068 |
| CT | Jun 2026 vs Jun 2025 | 1.13 | 0.98 | -0.1475 | -0.3167 | 0.0217 | 0.0876 |
| CT | May 2026 vs May 2025 | 1.04 | 0.97 | -0.0689 | -0.2253 | 0.0876 | 0.3882 |
| CT | Apr 2026 vs Apr 2025 | 0.98 | 0.98 | -0.0024 | -0.1514 | 0.1466 | 0.9747 |
| CT | Mar 2026 vs Mar 2025 | 1.08 | 1.02 | -0.0634 | -0.2362 | 0.1094 | 0.4721 |
| CT | Feb 2026 vs Feb 2025 | 1.06 | 1.14 | 0.0743 | -0.1096 | 0.2582 | 0.4286 |
| CT | Jan 2026 vs Jan 2026 | 1.07 | 1.01 | -0.0587 | -0.2195 | 0.1022 | 0.4749 |
| Non Obstetric Ultrasound | Jun 2026 vs Jun 2025 | 1.38 | 1.31 | -0.0681 | -0.2743 | 0.1382 | 0.5177 |
| Non Obstetric Ultrasound | May 2026 vs May 2025 | 1.33 | 1.33 | -0.0006 | -0.2006 | 0.1994 | 0.9954 |
| Non Obstetric Ultrasound | Apr 2026 vs Apr 2025 | 1.29 | 1.22 | -0.0723 | -0.2698 | 0.1253 | 0.4733 |
| Non Obstetric Ultrasound | Mar 2026 vs Mar 2025 | 1.4 | 1.34 | -0.0632 | -0.2563 | 0.1299 | 0.521 |
| Non Obstetric Ultrasound | Feb 2026 vs Feb 2025 | 1.35 | 1.33 | -0.0194 | -0.2011 | 0.1623 | 0.8341 |
| Non Obstetric Ultrasound | Jan 2026 vs Jan 2026 | 1.13 | 1.2 | 0.0704 | -0.1014 | 0.2421 | 0.4219 |
| Echocardiography | Jun 2026 vs Jun 2025 | 1.19 | 1.32 | 0.1335 | -0.0702 | 0.3372 | 0.1989 |
| Echocardiography | May 2026 vs May 2025 | 1.11 | 1.21 | 0.093 | -0.1255 | 0.3115 | 0.4042 |
| Echocardiography | Apr 2026 vs Apr 2025 | 1.07 | 1.18 | 0.1049 | -0.07 | 0.2798 | 0.2396 |
| Echocardiography | Mar 2026 vs Mar 2025 | 1.07 | 1.29 | 0.2139 | 0.0411 | 0.3867 | 0.0153 |
| Echocardiography | Feb 2026 vs Feb 2025 | 1.16 | 1.28 | 0.1131 | -0.0583 | 0.2846 | 0.1958 |
| Echocardiography | Jan 2026 vs Jan 2026 | 1.07 | 1.21 | 0.1447 | -0.012 | 0.3013 | 0.0703 |

**Appendix 7. Modelled proportions of patients waiting 6 weeks or longer for a diagnostic test, by local area, June 2025 versus June 2026**

*Preview: Full appendix available at* <https://github.com/rmjlrwh/DxWaitingTimes2>

| **Diagnostic test** | **Local area** | **Local area name** | **Observed % (waiting ≥6 weeks)Jun 2025** | **Observed lower CI — Jun 2025** | **Observed upper CI — Jun 2025** | **Observed count (waiting ≥6 weeks) — Jun 2025** | **Observed denominator (total waiting) — Jun 2025** | **Observed % (waiting ≥6 weeks)Jun 2026** | **Observed lower CI — Jun 2026** | **Observed upper CI — Jun 2026** | **Observed count (waiting ≥6 weeks) — Jun 2026** | **Observed denominator (total waiting) — Jun 2026** | **Modelled % (waiting ≥6 weeks) — Jun 2025** | **Modelled lower CI — Jun 2025** | **Modelled upper CI — Jun 2025** | **Modelled % (waiting ≥6 weeks) — Jun 2026** | **Modelled lower CI — Jun 2026** | **Modelled upper CI — Jun 2026** | **Absolute change (modelled)** | **Lower CI (change)** | **Upper CI (change)** | **P-value for time period comparison** |
| --- | --- | --- | --- | --- | --- | --- | --- | --- | --- | --- | --- | --- | --- | --- | --- | --- | --- | --- | --- | --- | --- | --- |
| Gastroscopy | 00L | NHS North East and North Cumbria ICB - 00L | 0.05 | 0.03 | 0.08 | 15 | 286 | 0.07 | 0.05 | 0.1 | 28 | 387 | 0.06 | 0.05 | 0.07 | 0.07 | 0.06 | 0.09 | 0.02 | 0.01 | 0.03 | 0 |
| Gastroscopy | 00N | NHS North East and North Cumbria ICB - 00N | 0.08 | 0.04 | 0.15 | 7 | 91 | 0.07 | 0.04 | 0.15 | 7 | 94 | 0.08 | 0.07 | 0.1 | 0.09 | 0.07 | 0.1 | 0 | -0.01 | 0.02 | 0.506 |
| Gastroscopy | 00P | NHS North East and North Cumbria ICB - 00P | 0.04 | 0.02 | 0.08 | 7 | 187 | 0.05 | 0.03 | 0.09 | 10 | 195 | 0.04 | 0.04 | 0.05 | 0.06 | 0.05 | 0.07 | 0.01 | 0.01 | 0.02 | 0.001 |
| Gastroscopy | 00Q | NHS Lancashire and South Cumbria ICB - 00Q | 0.16 | 0.12 | 0.21 | 49 | 303 | 0.05 | 0.03 | 0.08 | 14 | 283 | 0.16 | 0.13 | 0.18 | 0.06 | 0.05 | 0.07 | -0.1 | -0.12 | -0.08 | 0 |
| Gastroscopy | 00R | NHS Lancashire and South Cumbria ICB - 00R | 0.18 | 0.12 | 0.27 | 20 | 109 | 0.16 | 0.12 | 0.22 | 35 | 217 | 0.18 | 0.15 | 0.21 | 0.16 | 0.14 | 0.19 | -0.02 | -0.04 | 0.01 | 0.206 |
| Gastroscopy | 00T | NHS Greater Manchester ICB - 00T | 0.14 | 0.11 | 0.19 | 44 | 309 | 0.13 | 0.1 | 0.16 | 49 | 384 | 0.14 | 0.12 | 0.16 | 0.13 | 0.11 | 0.15 | -0.01 | -0.03 | 0.01 | 0.244 |
| Gastroscopy | 00V | NHS Greater Manchester ICB - 00V | 0.35 | 0.28 | 0.42 | 58 | 167 | 0.29 | 0.24 | 0.34 | 85 | 296 | 0.34 | 0.3 | 0.38 | 0.29 | 0.25 | 0.32 | -0.05 | -0.09 | -0.02 | 0.005 |
| Gastroscopy | 00X | NHS Lancashire and South Cumbria ICB - 00X | 0.67 | 0.62 | 0.72 | 239 | 357 | 0.14 | 0.1 | 0.2 | 25 | 173 | 0.66 | 0.62 | 0.7 | 0.16 | 0.14 | 0.18 | -0.5 | -0.53 | -0.46 | 0 |
| Gastroscopy | 00Y | NHS Greater Manchester ICB - 00Y | 0.3 | 0.24 | 0.37 | 56 | 187 | 0.31 | 0.27 | 0.36 | 132 | 424 | 0.3 | 0.26 | 0.33 | 0.31 | 0.27 | 0.35 | 0.01 | -0.02 | 0.05 | 0.419 |
| Gastroscopy | 01A | NHS Lancashire and South Cumbria ICB - 01A | 0.12 | 0.1 | 0.15 | 71 | 573 | 0.07 | 0.06 | 0.1 | 48 | 649 | 0.12 | 0.1 | 0.14 | 0.08 | 0.06 | 0.09 | -0.05 | -0.06 | -0.03 | 0 |
| Gastroscopy | 01D | NHS Greater Manchester ICB - 01D | 0.27 | 0.21 | 0.33 | 53 | 199 | 0.33 | 0.28 | 0.38 | 112 | 342 | 0.26 | 0.23 | 0.3 | 0.33 | 0.29 | 0.36 | 0.06 | 0.03 | 0.1 | 0.001 |
| Gastroscopy | 01E | NHS Lancashire and South Cumbria ICB - 01E | 0.65 | 0.6 | 0.69 | 293 | 452 | 0.14 | 0.1 | 0.19 | 28 | 202 | 0.64 | 0.6 | 0.68 | 0.15 | 0.13 | 0.18 | -0.49 | -0.52 | -0.45 | 0 |
| Gastroscopy | 01F | NHS Cheshire and Merseyside ICB - 01F | 0.05 | 0.02 | 0.1 | 5 | 110 | 0.15 | 0.11 | 0.2 | 36 | 247 | 0.06 | 0.05 | 0.07 | 0.14 | 0.12 | 0.17 | 0.08 | 0.07 | 0.1 | 0 |

**Appendix 8. Summary of relative change in modelled proportions of patients waiting 6 weeks or longer for a diagnostic test (number and proportion of local areas), June 2025 versus June 2026**

| **Test** | **≥75% reduction (n)** | **≥75% reduction (%)** | **50–75% reduction (n)** | **50–75% reduction (%)** | **0–50% reduction (n)** | **0–50% reduction (%)** | **No significant difference (n)** | **No significant difference (%)** | **0–50% increase (n)** | **0–50% increase (%)** | **50–100% increase (n)** | **50–100% increase (%)** | **≥100% increase (n)** | **≥100% increase (%)** |
| --- | --- | --- | --- | --- | --- | --- | --- | --- | --- | --- | --- | --- | --- | --- |
| Gastroscopy | 2 | 1.89 | 3 | 2.83 | 23 | 21.7 | 23 | 21.7 | 20 | 18.87 | 11 | 10.38 | 21 | 19.81 |
| Colonoscopy | 2 | 1.89 | 8 | 7.55 | 19 | 17.92 | 18 | 16.98 | 16 | 15.09 | 17 | 16.04 | 23 | 21.7 |
| Flexible Sigmoidoscopy | 3 | 2.83 | 7 | 6.6 | 23 | 21.7 | 16 | 15.09 | 19 | 17.92 | 15 | 14.15 | 20 | 18.87 |
| Cystoscopy | 1 | 0.94 | 12 | 11.32 | 30 | 28.3 | 19 | 17.92 | 24 | 22.64 | 7 | 6.6 | 10 | 9.43 |
| MRI | 2 | 1.89 | 11 | 10.38 | 21 | 19.81 | 16 | 15.09 | 13 | 12.26 | 9 | 8.49 | 31 | 29.25 |
| CT | 6 | 5.66 | 16 | 15.09 | 20 | 18.87 | 19 | 17.92 | 12 | 11.32 | 11 | 10.38 | 19 | 17.92 |
| Non Obstetric Ultrasound | 16 | 15.09 | 11 | 10.38 | 20 | 18.87 | 18 | 16.98 | 4 | 3.77 | 6 | 5.66 | 28 | 26.42 |
| Echocardiography | 9 | 8.49 | 18 | 16.98 | 14 | 13.21 | 17 | 16.04 | 14 | 13.21 | 9 | 8.49 | 22 | 20.75 |

**Appendix 9. Modelled and observed proportion of patients waiting 6 weeks or longer in June 2025 versus June 2026, for individual local areas**

Ranked by proportion in June 2025. Dots = predicted; crosses = observed

**a) Endoscopy**

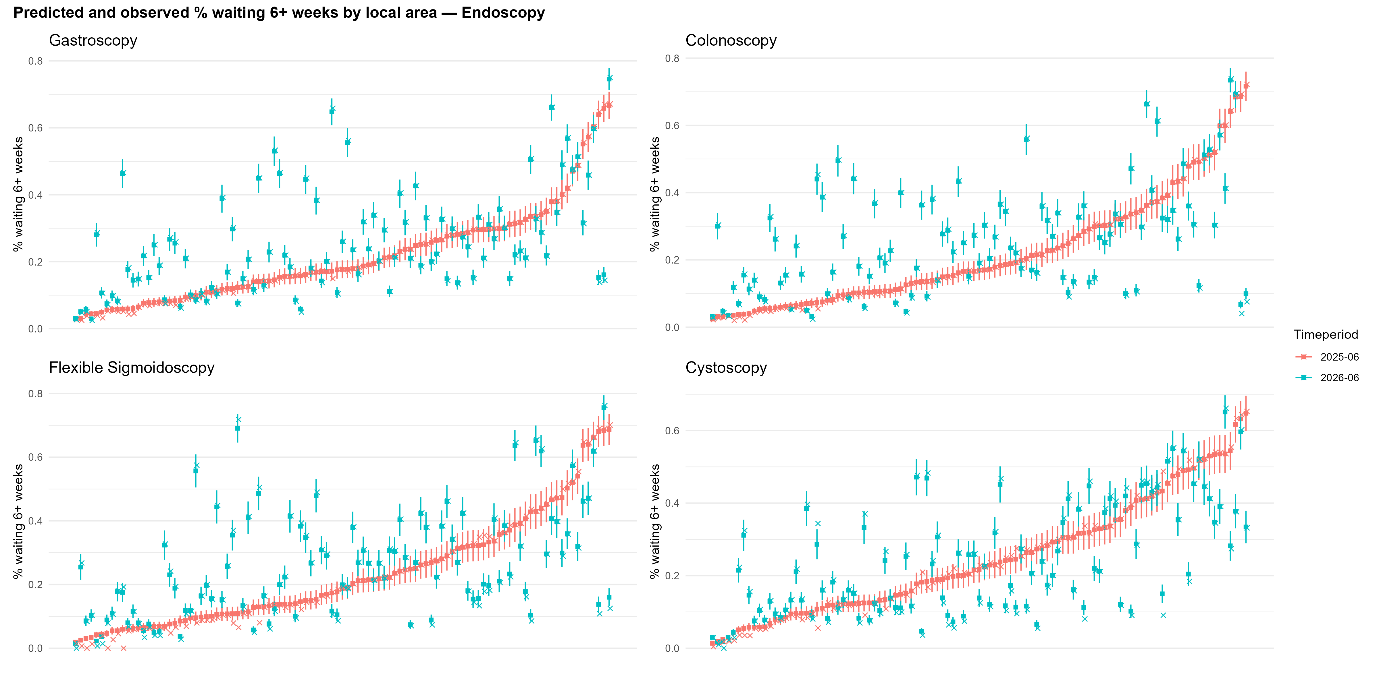

**b) Imaging**

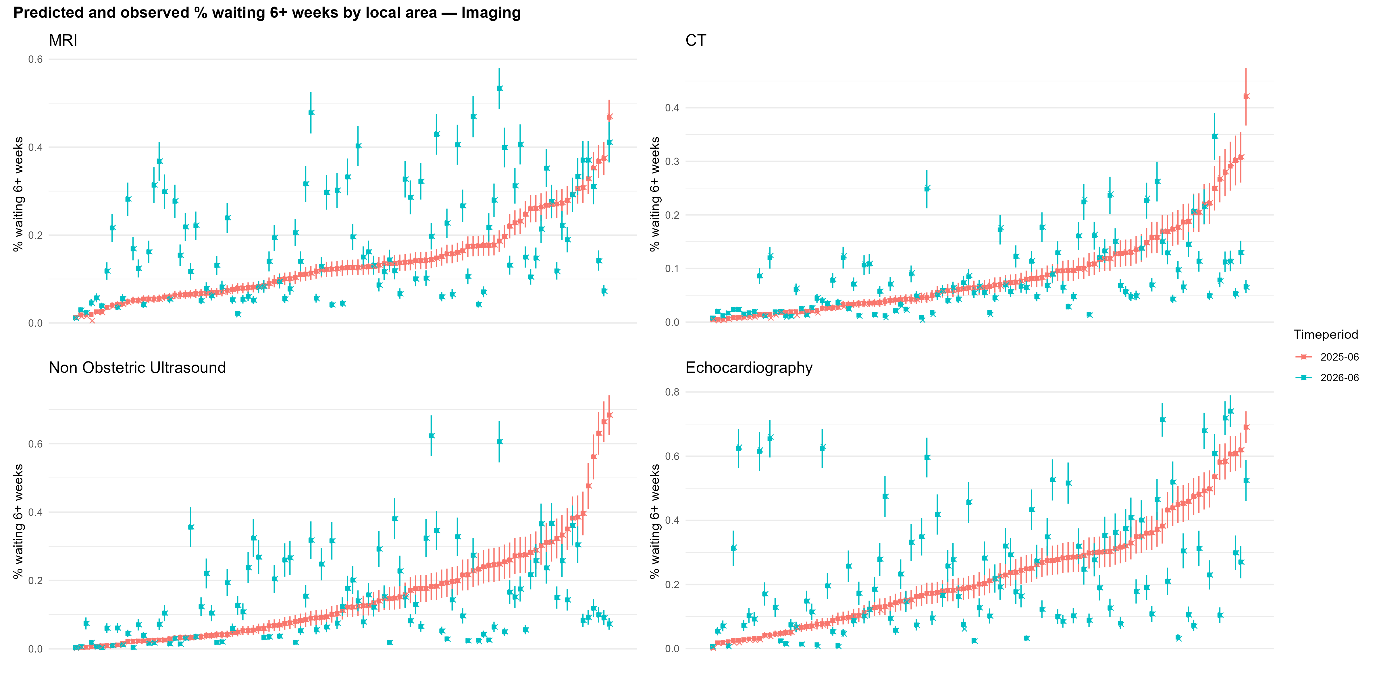
